# LiteRev, an Automation Tool to Support Literature Reviews: A Case Study on Acute and Early HIV Infection in Sub-Saharan Africa

**DOI:** 10.1101/2023.02.20.23286179

**Authors:** Erol Orel, Iza Ciglenecki, Amaury Thiabaud, Alexander Temerev, Alexandra Calmy, Olivia Keiser, Aziza Merzouki

## Abstract

**Background:** Literature Reviews (LRs) identify, evaluate, and synthesise relevant papers to a particular research question to advance understanding and support decision making. However, LRs, especially traditional systematic reviews are slow, resource intensive, and are outdated quickly.

**Objective:** Using recent Natural Language Processing (NLP) and Unsupervised Machine Learning (UML) methods, this paper presents a tool named LiteRev that supports researchers in conducting LRs.

**Methods:** Based on the user’s query, LiteRev can perform an automated search on different open-access databases and retrieve relevant metadata on the resulting papers. Papers (abstracts or full texts) are text processed and represented as a Term Frequency-Inverse Document Frequency (TF-IDF) matrix. Using dimensionality reduction (PaCMAP) and clustering (HDBSCAN) techniques, the corpus is divided into different topics described by a list of keywords. The user can select one or several topics of interest, enter additional keywords to refine their search, or provide key papers to the research question. Based on these inputs, LiteRev performs an iterative nearest neighbours search, and suggests a list of potentially interesting papers. The user can tag the relevant ones and trigger a new search until no additional paper is suggested for screening. To assess the performance of LiteRev, we ran it in parallel to a manual LR on the burden and care for acute and early HIV infection in sub-Saharan Africa. We assessed the performance of LiteRev using True and False Predictive Values, recall and Work Saved over Sampling.

**Results:** We extracted, text processed and represented into a TF-IDF matrix 631 unique papers from PubMed. The topic modelling module identified 5 main topics and 16 topics (ranging from 13 to 98 papers) and extracted the 10 most important keywords for each. Then, based on 18 key papers, we were able to identify 2 topics of interest with 7 key papers in each of them. Finally, we ran the k-nearest neighbours module and LiteRev suggested first a list of 110 papers for screening, among which 45 papers were confirmed as relevant. From these 45 papers, LiteRev suggested 26 additional papers, out of which 8 were confirmed as relevant. At the end of the iterative process (4 iterations), 193 papers out of 613 papers in total (31.5% of the whole corpus) were suggested by LiteRev. After title/abstract screening, LiteRev identified 64 out of the 87 relevant papers (i.e., recall of 73.6%). After full text screening, LiteRev identified 42 out of the 48 relevant papers (i.e., recall of 87.5%, and Work Saved over Sampling of 56.0%).

**Conclusions:** We presented LiteRev, an automation tool that uses NLP and UML methods to streamline and accelerate LRs and to support researchers in getting quick and in-depth overviews on any topic of interest.

## Introduction

Recently, the traditional emphasis of Literature Reviews (LRs) in identifying, evaluating, and synthesising all relevant papers to a particular research question has shifted towards mapping research activity and consolidating existing knowledge [1]. Despite this broader scope, manual LRs are still error-prone, time and resource-intensive and have become ever more challenging over the years due to the increasing number of papers published in academic databases. It is estimated that within two years of publication, about one fourth of all LRs are outdated, as reviewers fail to incorporate new papers on their topic of interest [2,3].

To shorten time to completion, automation tools have been developed to either fully automate or semi-automate one or more specific tasks involved in conducting a LR, such as screening titles and abstracts [4,5], sourcing full texts or automating data extraction [6]. Also, recent advances in Natural Language Processing (NLP) and Unsupervised Machine Learning (UML) have produced new techniques that can accurately mimic manual LRs faster and at lower costs [7,8,9]. In Vienna, in 2015, the International Collaboration for the Automation of Systematic Reviews (ICASR) was initiated to establish a set of principles to enable tools to be developed and integrated into toolkits [10]. Also, since 2021, an open source machine learning framework named ASReview is under development for efficient and transparent systematic reviews [11].

In 2020, our group of researchers started developing an automation tool for LRs [12] in order to obtain a comprehensive overview of the sociobehavioral factors influencing HIV prevalence and incidence in Malawi. In this paper, we propose LiteRev, a new version of our automation tool that overcomes some of the shortcomings of our previous tool. While previously restricted to Paperity, PubMed, PubmedCentral, JSTOR, and arXiV, the search now includes two primary preprint services in the field of epidemiology and medical sciences, biorXiV and medrXiV, and CORE, a large collection of open-access research papers. Also in our previous tool, the search was systematically performed on the papers’ full texts and references were included in the processed text. In LiteRev, the user can choose to focus on the abstract or on the full text and include or exclude the references. In addition, multiple parallel Application Programming Interfaces (APIs) connections to each database have been implemented, allowing for faster retrieval of papers. Since two years, NLP and UML have rapidly evolved, and LiteRev makes use of the most recent text processing, embedding and clustering techniques. Finally, we added a k-nearest neighbour’s search module that allows the user to find papers of high similarities with key papers to the research question.

In order to assess the performance of LiteRev, we conducted a manual LR using one open-access database, PubMed, and two subscription-based databases, Embase and Web of Science. All papers available by the 20th of December 2022 and related to burden and care for acute and early HIV infection in sub-Saharan Africa were retrieved and after removing duplicates, unique papers were screened for relevance. After screening, papers from PubMed identified as relevant by the manual LR were compared to the list of suggested papers by LiteRev. We discussed the performance using standard classification metrics such as True and False Predictive Values, recall, and Work Saved over Sampling (WSS).

## Methods

### LiteRev

#### Metadata collection and text processing

Based on the user’s query, LiteRev can perform an automated search, using the corresponding APIs, on 8 different open-access databases: PubMed, PubMedCentral, CORE, JSTOR, Paperity, arXiv, biorXiv and medXriv. Available metadata, i.e., list of authors and their affiliations, MesH keywords, Digital Object Identifier (DOI), title, abstract, publication date, journal provider, and URL of the PDF version of the full text paper, are retrieved and stored in a PostgreSQL database. If the full text is not available as a metadata, it is extracted automatically from the available PDF file, then, references, acknowledgements, and other unnecessary terms are removed and the remaining text is checked to confirm that it still satisfies the search terms. Duplicated papers are merged, collecting as much information as possible on the same paper from different sources. Depending on the user’s needs and requirements, LiteRev can be performed on the full text or on the abstract.

Natural Language Processing has evolved rapidly, and, in particular, some powerful tools were developed to process text data much more efficiently. We included those features into LiteRev (Gensim [13] and Spacy [14]). After removing papers with empty text, emails, newline characters, single quotes, internet addresses, and punctuation are stripped and papers that do not fulfil the language(s) (one or multiple) chosen by the user are discarded. Sentences are then split into words and lemmatised to remove as many variations of the same word as possible. Words belonging to a list of stop-words (i.e., words that are not informative) and words with less than three characters are also removed. Next, bigrams, trigrams and four-grams (i.e., the combination of two, three and four words) are created using a probabilistic measure. In practice, n-gram models are highly effective in modelling language data. Finally, we remove words that are in only one paper or words that occur too often (i.e., in more than 80% of the corpus) to have a significant meaning.

#### Clustering and topic modelling

Topic modelling allows organising documents into clusters based on similarity, and identifying abstract topics covered by similar papers. In LiteRev, it allows the user to broaden the search strategy and to get a more comprehensive and organised overview of the corpus. It can also help to quickly discard a pool of papers when searching the literature for a specific topic and significantly reduce the amount of text to verify manually.

After abstracts or full texts are processed, each paper’s remaining words (namely bags of words) are represented as a Term Frequency-Inverse Document Frequency (TF-IDF) matrix which is computed using the Scikit-Learn package [15]. A TF-IDF matrix is similar to a document (in row) - word (in column) co-occurrence matrix, normalised by the number of papers in which the word is present. Less meaningful words, often present in the corpus, get a lower score. Because of the often-high dimension of the TF-IDF matrix (size of corpus x size of vocabulary), it is needed to embed the matrix using a Pairwise Controlled Manifold Approximation (PaCMAP) dimensionality reduction technique [16]. The corpus is then divided into different clusters using the Hierarchical Density-Based Spatial Clustering of Applications with Noise (HDBSCAN) algorithm [17]. While HDBSCAN allows to classify some papers as noise, we have decided to consider those as a cluster in itself.

PaCMAP and HDBSCAN have several important hyperparameters that need to be determined. Table S1 in the Supplementary Material represents the 4 hyperparameters involved and the ranges of their possible values. To find the best set of hyperparameters possible, we use the Tree-structured Parzen Estimator algorithm implemented by the Optuna package [18] and store the results of 500 trials in the previously created PostgreSQL database. The Density-Based Clustering Validation (DBCV), a weighted sum of “Validity Index” values of clusters [19], is the considered performance metric to compare the different sets. Its value varies between 0 and 1 when used with HDBSCAN, with larger values providing better clustering solutions. This metric takes the noise into account and captures the shape property of clusters via densities and not distances. For coherency check, another metric is computed, the Silhouette coefficient, which measures cluster cohesiveness and separation with an index between -1 to 1, with larger values providing better clustering solutions [20].

If after 500 trials, the DBCV score is below 0.5, another round of 500 trials is performed, and so on until a DBCV score equal or above 0.5 is reached. Once the values of the hyperparameters that maximise the DBCV score are determined, obtained clusters that are larger than 25% of the corpus are clustered again with the same entire procedure described above (starting from the text processing). Once each cluster is smaller than 25% of the corpus, its 10 most important words are extracted using the YAKE package to ensure interpretability and define topics. This supports the user in getting a quick overview of the corpus and, if desired, to select one or more topics of interest for further exploration. They can then also enter additional keywords to refine his search.

#### Nearest neighbours

LiteRev allows the user to define papers in the corpus as being key to the research question (or to add them). Using the k-nearest neighbour (k-NN) algorithm from the Scikit-Learn package [15], a list of potentially relevant papers is provided to the user. Papers deemed to be relevant are tagged by the user and considered as new key papers. This process is iterated as long as relevant papers are being identified (generally 3 to 4 iterations). The initial value of the hyperparameter k, which represents the number of nearest neighbours to be selected, is equal to the value of the number of neighbours for PaCMAP obtained at the first clustering process. The dimension space is the same as the number of dimensions obtained during the embedding process by PaCMAP.

The list of relevant papers from the k-NN search and/or a list of papers about one or more topics can then be exported in a csv or html format and their pdf retrieved and stored in a zip folder. For visualisation and further exploration, an interactive 2D representation of the corpus is available in a html format. Every dot, coloured according to the cluster it belongs to, represents a paper with the following available information: date, title, 10 most important keywords of the cluster’s topic and the cluster number. When clicking on a paper (dot), a direct access to the full text is provided using the URL. The diagram in Figure 1 shows the entire process flow of LiteRev.

**Figure 1.**
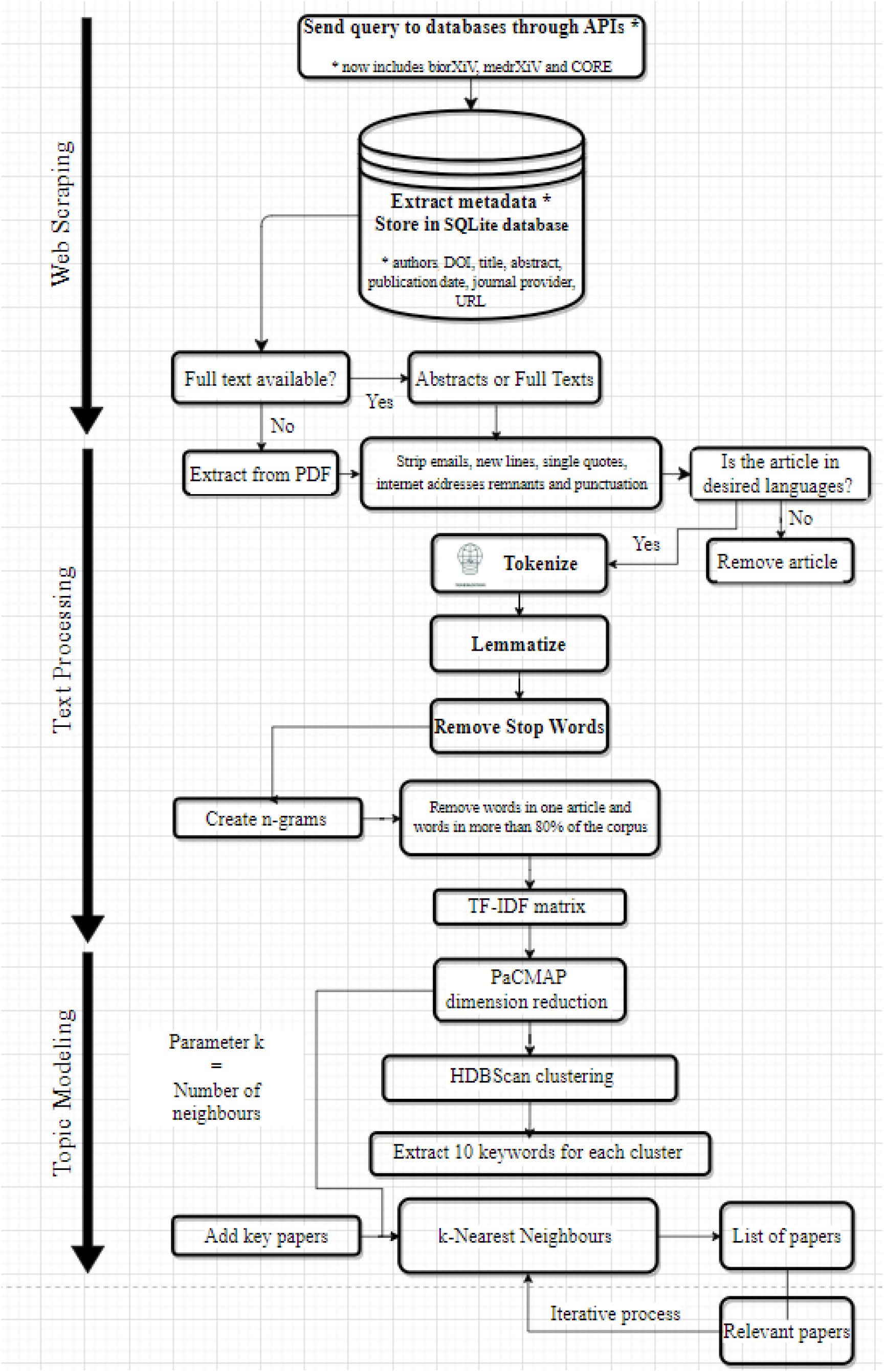
Diagram of LiteRev process

### Manual Literature Review

The manual LR aimed at summarising the current evidence on burden and care provided for acute and early HIV infection (AEHI) in sub-Saharan Africa, to inform policy, practice and research in future, addressing the following questions:

- What is the prevalence of AEHI in sub-Saharan Africa among people being tested for HIV?
- What models of care have been used for AEHI diagnosis and care, including treatment, partner notifications and behaviour change?
- What linkage to care has been reached?
- What facilitators and barriers to AEHI care were identified?

We searched all papers in PubMed, Embase and Web of Science related to burden and care for acute and early HIV infection in sub-Saharan Africa that were published from the inception of the databases to December 20 2022, using the query: “(“early hiv” OR “primary hiv” OR “acute hiv” OR “HIV Human immuno deficiency virus” OR (“Window period” AND HIV)) AND (“Africa South of the Sahara” OR “Central Africa” OR “Eastern Africa” OR “Southern Africa” OR “Western Africa” OR “sub-saharan africa” OR “subsaharan africa” OR angola OR benini OR botswana OR “burkina faso” OR burundi OR cameroon OR “cape verde” OR “central africa” OR “central african republic” OR chad OR comoros OR congo OR “cote d ivoire” OR “democratic republic congo” OR djibouti OR “equatorial guinea” OR eritrea OR eswatini OR ethiopia OR gabon OR gambia OR ghana OR guinea OR “guinea-bissau” OR kenya OR lesotho OR liberia OR madagascar OR malawi OR mali OR mayotte OR mozambique OR namibia OR niger OR nigeria OR rwanda OR sahel OR “sao tome and principe” OR senegal OR “sierra leone” OR somalia OR “south africa” OR “south sudan” OR sudan OR tanzania OR togo OR uganda OR zambia OR zimbabwe)”. This query is specific to PubMed syntax and is the exact same both for the manual LR and for LiteRev. Syntax specific queries for the manual LR in Embase and Web of Science are to be found in the Supplementary Material. Papers retrieved from Embase and Web of Science have not been used by LiteRev and will not be part of the comparison and performance assessment but its results will be discussed in the Results and Discussion section.

The studies were included if they described AEHI prevalence among population tested for HIV and/or describe the diagnostic strategy, model of care and/or linkage to care for AEHI, including studies looking at perceptions and barriers among patients and staff. Only studies conducted in sub-Saharan Africa were included. We followed the JBI methodology for conducting LRs [22] and papers identified by the databases were uploaded into Rayyan [23]. Duplicates were deleted and the screening process, on titles and abstracts, was conducted independently by 2 reviewers (EO and IC). Selected papers were further manually screened based on full text for eligibility against inclusion criteria. LiteRev was run in parallel on the abstracts only but results were compared both to the title/abstract screening phase and to the full text screening phase of the manual LR.

### Performance Comparison

In order to assess the performance of LiteRev, we compared the results from the manual LR to the same review conducted using LiteRev. Relevant and not relevant papers, as identified by the manual LR during the title/abstract screening phase and the full text screening phase, were defined as true labels. Suggested and not suggested papers by LiteRev were considered as predicted labels. Based on these figures, two confusion matrices were produced. Positive and Negative Predictive Values (% of relevant and not relevant papers correctly identified; PPV and NPV), recall (number of relevant papers identified using LiteRev among those identified using manual review) and Work Saved over Sampling (WSS) [24,25], percentage of abstracts or full texts that the user did not have to read because they were not suggested for screening by LiteRev, were computed and discussed.

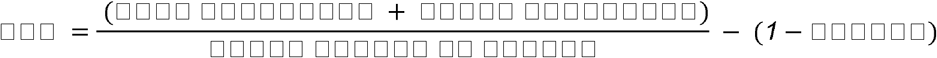

Where

- True negatives is the number of non-relevant abstracts that were correctly identified as non-relevant by LiteRev, i.e. that were not suggested by LiteRev for screening,
- False negatives is the number of relevant abstracts incorrectly classified as non-relevant by LiteRev.

## Results

### LiteRev

#### Text Processing and Topic Modelling

Based on the search strategy described in the Methods section, we obtained 653 papers with metadata directly from PubMed and added one key paper given by the user that was not present in the list of retrieved papers. After removing duplicates (3), papers with no abstract available (15), those not in english (3), and empty abstract after text processing (2), 631 unique papers were transformed in a TF-IDF matrix comprised of 631 rows representing the corpus and 3’136 columns representing the unique words (vocabulary), including n-grams.

For the first embedding and clustering process, a DBCV score of 0.533 was obtained after the first 500 trials with the following best set of hyperparameters: PaCMAP: 310 dimensions and 18 neighbours; HDBSCAN: minimum cluster size of 30 and minimum samples of 7. This resulted in 5 main clusters composed of respectively 203, 193, 169, 35 and 31 papers. The 3 largest main clusters contained more than 25% of the total number of papers in the corpus, which triggered 3 additional text processing, embedding and clustering processes. The best set of hyperparameters for these additional processes can be found in Table S1 in the Supplementary Material.

At the end, the pool of 203 papers were splitted in 5 clusters (with respectively 98, 41, 25, 21 and 18 papers), the pool of 193 papers in 7 clusters (with respectively 47, 40, 37, 22, 20 14 and 13 papers) and the pool of 169 papers in 2 clusters (with respectively 87 and 82 papers). In total, the corpus of 631 papers was divided in 16 clusters ranging from 13 to 98 papers. Figure 3 shows the 2D map of the corpus with the 16 clusters identified. Table 1 shows the corresponding 16 topics grouped by main topics, described by their 10 most important keywords and the number of papers in each.

**Table 1.** The 16 topics grouped by main topics (in blue) with the 10 most important keywords, the number of papers and the number of relevant papers in total (key papers)

| Topic | Keywords | # of papers | # of relevant papers (key) |
| --- | --- | --- | --- |
| **woman, patient, risk, year, treatment, associate, month, incidence, testing, care** |  |  |  |
| 0 | woman, risk, year, incidence, high, man, partner, transmission, sexual, testing | 98 | 9 (1) |
| 1 | cart, month, initiation, group, treatment, viral, rna, child, infant, week | 18 | 0 (0) |
| 2 | risk, high, health, day, score, aehi, prevalence, care, diagnosis, population | 41 | 6 (2) |
| 3 | patient, treatment, care, late, diagnosis, associate, testing, aor, datum, initiation | 21 | 0 (0) |
| 4 | patient, disease, adult, infect, lymphadenopathy, cell, tuberculous, lymphadenitis, associate, present | 25 | 0 (0) |
| **cell, viral, response, virus, subtype, individual, primary, isolate, antibody, infect** |  |  |  |
| 5 | antibody, response, neutralize, vaccine, isolate, neutralization, epitope, env, primary, individual | 47 | 0 (0) |
| 6 | subtype, resistance, drug, sequence, mutation, diversity, strain, primary, patient, recombinant | 40 | 3 (0) |
| 7 | response, specific, associate, increase, immune, ifn, early, gag, point, level | 37 | 0 (0) |
| 8 | level, viremia, acute, associate, early, individual, infect, load, cytokine, set | 20 | 1 (0) |
| 9 | load, early, copy, log, plasma, subtype, woman, time, african, rna | 22 | 5 (1) |
| 10 | isolate, primary, tropic, individual, derive, clone, strain, infect, dual, sequence | 13 | 0 (0) |
| 11 | response, immune, phi, specific, control, activation, plasma, individual, acute, cytokine | 14 | 0 (0) |
| **test, testing, blood, ahi, risk, acute, positive, care, sample, assay** |  |  |  |
| 12 | blood, assay, sample, donor, positive, risk, incidence, antibody, estimate, acute | 82 | 21 (7) |
| 13 | ahi, care, participant, health, intervention, patient, diagnosis, early, acute, risk | 87 | 37 (7) |
| **infant, mother, week, transmission, child, month, age, woman, test, infect** |  |  |  |
| 14 | infant, mother, week, transmission, child, month, age, woman, test, infect | 35 | 0 (0) |
| **child, year, mortality, age, infect, treatment, patient, associate, month, clinical** |  |  |  |
| 15 | child, year, mortality, age, infect, treatment, patient, associate, month, clinical | 31 | 0 (0) |

**Figure 2.**
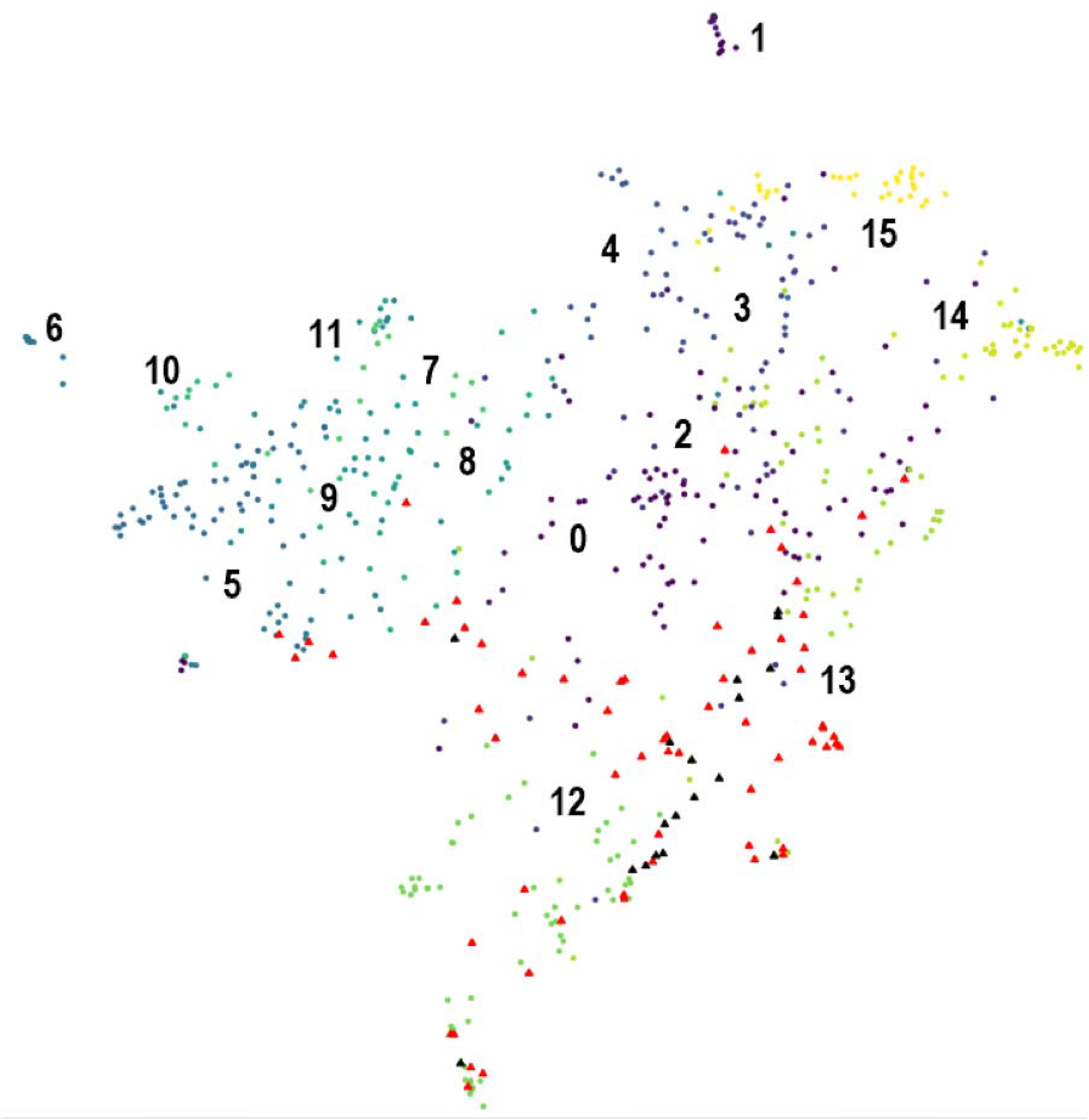
2D representation of the corpus with the 16 clusters. Black triangles represent the 18 key papers and red triangles represent the 64 relevant papers correctly identified by LiteRev

**Figure 3.**
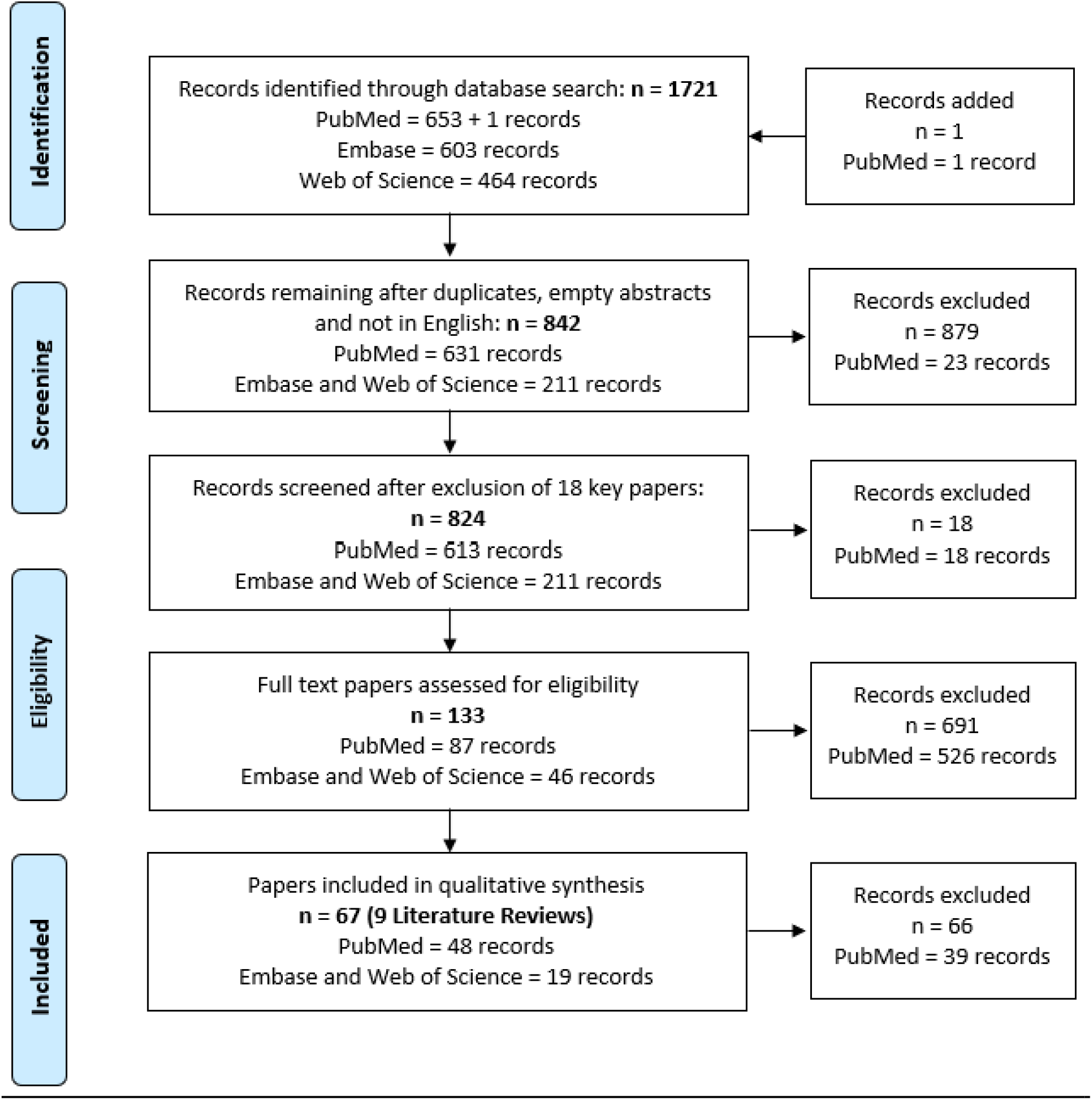
PRISMA diagram of the manual LR related to burden and care for acute and early HIV infection in sub-Saharan Africa

### Manual Literature Review

Using the search query described in the Methods section, 1’721 records were retrieved, among which 653 were from PubMed and 1’067 records from 2 subscription-based databases, namely Embase and Web of Science. 879 records were excluded after removing duplicates, empty abstracts and papers that were not written in English. This resulted in 631 unique papers in PubMed and 211 unique papers in Embase and Web of Science. We also removed the 18 key papers out of the PubMed corpus before the screening phases. In total, 613 papers in PubMed were screened at the title and abstract level and 87 of them were relevant to the research question. After the full text screening phase on these 87 relevant papers, we found 48 papers to be relevant with the manual LR.

Out of the 211 unique papers from Embase and Web of Science, 46 papers were found relevant to the research question after the title/abstract screening phase (i.e., 34.6% of all relevant papers), and 19 after the full text screening phase (i.e., 28.4% of all relevant papers) (Figure 3). From these 19 relevant papers, 3 were conference abstracts and 1 paper was kept only based on its title and abstract as the full text couldn’t be found. These 221 papers were not part of PubMed, and hence, not available to LiteRev.

### Nearest Neighbours Search and Performance Comparison

We were provided by the user (IC) a list of 18 key papers on the topic. With these 18 papers, we performed a k-nearest neighbours search on the corpus, embedded into 310 dimensions, with k=18, the number of the nearest neighbours for PaCMAP that maximised the DBCV score of the first clustering process. The first k-NN search suggested 110 papers, including 45 of the relevant papers identified by the manual LR title/abstract screening (precision of 41%). Based on these 45 relevant papers, the second k-NN iteration suggested 26 additional papers out of which 8 were confirmed as relevant (precision of 31%). The third iteration found 9 more relevant papers out of 38 papers suggested (precision of 24%). The fourth and last iteration suggested 19 papers out of which 1 was relevant (precision of 5%).

In total, 193 papers out of the 613 papers were suggested by LiteRev. Suggested papers included 64 of the 87 papers identified as relevant during the title/abstract screening of the manual LR. Figure 3 maps the key papers (black triangles) and the relevant papers (red triangles) identified at the title/abstract screening level of the manual LR, and that were correctly classified as relevant by LiteRev. Table 1 indicates the number of key papers and the number of relevant papers in each topic.

Figure 4 (top panel) summarises the above results and represents the confusion matrix between LiteRev (Predicted labels) and the manual LR (True labels) after the title/abstract screening phase. Based on these numbers, the PPV was 33.2%, the NPV was 94.5% and the recall was 73.6%, which led to a WSS of 42.1%.

**Figure 4.**
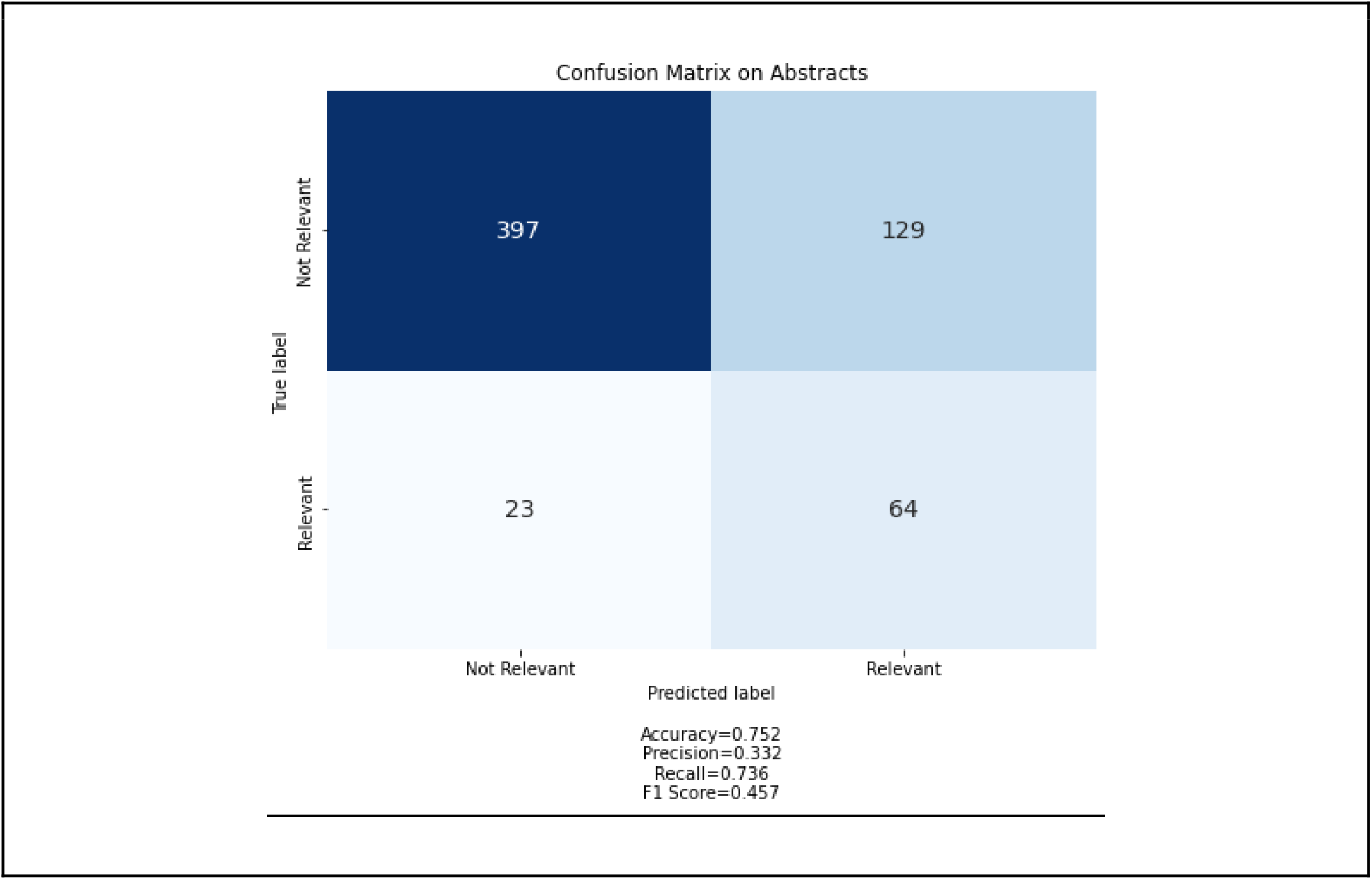
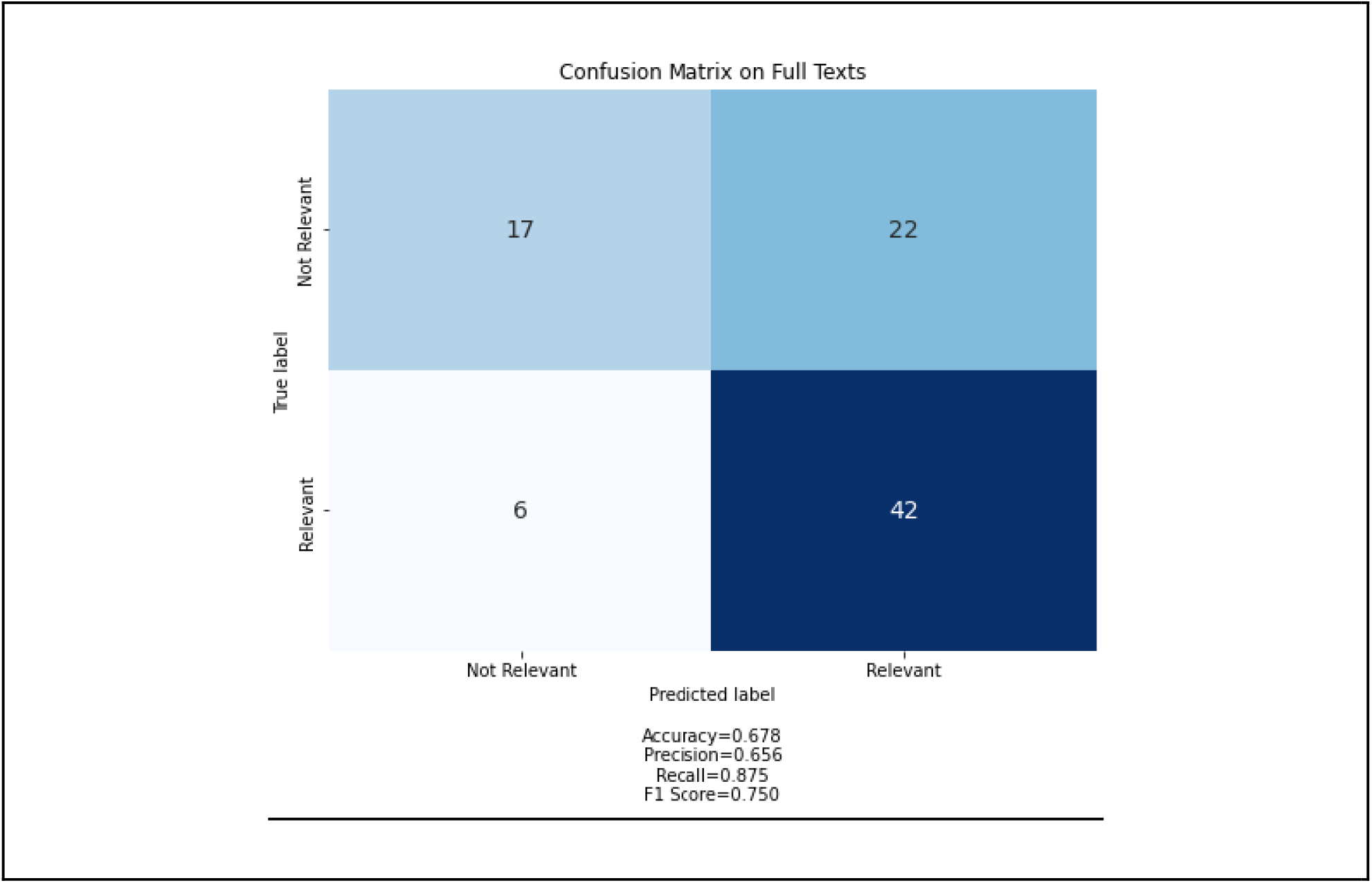
Confusion Matrices based on the results of (top panel) the title/abstract screening, and (bottom panel) full-text screening performed during the manual LR.

The 64 relevant papers found by LiteRev belonged essentially to 2 topics (30 releva papers in one and 14 relevant papers in the other). The topic that contained 30 relevant papers had 87 papers in total, and covered early diagnosis, care seeking and interventions during Acute HIV Infection stage (keywords: “ahi, care, participant, health, intervention, patient, diagnosis, early, acute, risk”). The topic that contained 14 relevant papers had 82 papers, and covered the detection of AHI by antibody assays and incidence estimate (keywords: “blood, assay, sample, donor, positive, risk, incidence, antibody, estimate, acute”). Screening 53 additional papers (those not suggested by the nearest neighbours search) from these 2 topics would allow the user to identify 3 additional relevant papers.

After the full text screening phase of the manual LR, 48 out the 87 relevant papers from the title and abstract screening phase were deemed relevant to the research question. The list of (64) relevant papers suggested by LiteRev (based on abstracts only), included 42 out of the 48 papers confirmed as relevant after the full text screening phase of the manual LR. Figure 4 (bottom panel) summarises the above results and represents the confusion matrix between LiteRev (Predicted labels) and the manual LR (True labels) after the full text screening phase. Based on these numbers, the PPV was 65.6%, the NPV was 26.1% and the recall was 87.5%, which led to an additional WSS of 13.9% for an overall WSS of 56.0% compared to the manual LR.

#### Processing time

The processing time represents the overall computation time taken by LiteRev to complete the entire process of metadata retrieval, processing, clustering, and neighbour search. It doesn’t include the time that the user took to check the relevance of the suggested papers. The percentage of time saved by the user, is expressed by the work saved over sampling (WSS) metric.

It took 5 minutes for LiteRev to retrieve the metadata of the 653 papers and to text process the remaining 631 abstracts and transform it into a TF-IDF matrix. It took an additional two days for the main clustering and the 3 additional clustering processes. Each trial of the optimization process with a specific set of hyperparameters required on average 1 minute of computation. With 3’000 trials in total (500 for the main clustering process, 1’000 for the first two additional clustering processes and 500 for the last one) run sequentially, this led to an additional 50 hours, i.e., roughly 2 days, to complete the entire optimisation process. This computation time can be substantially reduced by running the trials in parallel. Finally, the nearest neighbours are obtained almost instantaneously.

## Discussion

### Principal Results

We presented LiteRev, an automation tool that uses NLP and UML methods to support researchers in different steps of a manual LR. The identification of papers to be included in a LR is a critical and time-intensive process, with the majority of time spent in screening thousands of papers for relevance. By combining text processing, literature mapping, topic modelling and similarity-based search, LiteRev provides a fast and efficient way to remove duplicates, select papers from specific languages, visualise the corpus on a 2D map, identify the different topics covered when addressing the research question and suggest a list of potentially relevant papers to the user based on their input (e.g., prior knowledge of key papers).

Preliminary usage of LiteRev showed that it significantly reduced the researcher’s workload and overall time required to perform a LR. Compared to a manual LR, LiteRev correctly identified 87.5% of relevant papers (recall), by screening only 31.5% of the whole corpus, which corresponds to a total Work Saved over Sampling of 56.0% (WSS) at the end of the full text screening phase. In addition, the actual time spent on running LiteRev and retrieving the results was relatively short, and the user was free in the meantime to focus on other work. The text processing and the nearest neighbours search took no more than 5 minutes of computation for 631 papers.

With its topic modelling capability, LiteRev aims at summarising current evidence on a specific research question, to inform policy, practice and research. For our use case, LiteRev identified 5 main topics and 16 different topics related to acute and early HIV infection in Sub-Saharan Africa, allowing the researcher to have an overview on the different perspectives related to this research question. Finding 61 out of the 105 relevant papers after the title and abstract screening phase (including the key papers) in only 2 topics validates the quality of the clustering.

### Limitations

LiteRev is currently limited to open-access databases that provide free APIs to abstract or full text papers. Databases often used for LRs, such as Embase or Web of Science do not provide APIs access, require a subscription for accessing full text papers or do not allow for text mining and machine learning analysis. Hence, 19 relevant papers identified in Embase or Web of Science were not available to LiteRev. Also, when performed on full texts, LiteRev currently works on digitally-generated PDFs, but not on image-only (scanned) PDFs.

Another limitation concerns the possibility of sharing the list of potentially relevant papers with other users/reviewers. LiteRev does not offer this functionality yet, hence double screening of papers and comparison of results is not possible at the moment. To overcome this limitation, the user has the option to export its list of papers into a csv format uploadable on Rayyan or other similar softwares for systematic reviews.

As of today, LiteRev is still intended to complement rather than replace full systematic reviews. Finally, by January 2023, no public web-based User Interface (UI) is available yet.

### Future work

O’Connor et al. [26] found that overall, many of the automation tools were not compatible with current practice, because they were not easily integrated into current workflows and not particularly easy to use for nontechnical persons. Also, there was not enough evidence of accuracy to earn the trust of reviewers. LiteRev is developed in an iterative and interactive way by continuously integrating feedback from users and its modules can easily be updated or replaced depending on the needs of the users and the technical evolutions. We are further developing LiteRev by proposing a web application with a user-friendly interface and by adding more functionality in order to better automate the different stages of a LR. We are also planning to implement a living review [27] by retrieving new papers on each research questions in our database (e.g., “HIV” AND “Africa”) on a regular basis (e.g., every month) and each new paper will be text processed and assigned to the topic it belongs to using a predictive algorithm. Although we compared the performance of LiteRev with one manual LR in this paper, we plan to perform additional similar comparisons and performance evaluations in the future, using other published LRs covering different topics.

## Data Availability

All data produced in the present study are available upon reasonable request to the authors.

## Conclusions

We presented LiteRev, an automation tool that uses NLP and UML techniques to support, facilitate and accelerate the conduction of Literature Reviews providing aid and automation to different steps involved in this process. Its different modules (retrieval of papers’ metadata from open-access databases using a search query, processing of texts, embedding and clustering, and finding of nearest neighbours) can easily be updated or replaced depending on the needs of the users and the technical evolutions. As more papers are published every year, LiteRev not only has the potential to simplify and accelerate LRs, but it also has the capability of helping the researcher get a quick and in-depth overview on any topic of interest.

### Financial disclosure

We acknowledge the support of the Swiss National Science Foundation (SNF professorship grants n°196270 and n°202660 to Prof. O. Keiser), which funded this study. The funder had no role in study design, data collection and analysis, decision to publish, or manuscript preparation.

## Authors’ Contributions

EO and ATh wrote the code in Python. EO and AM obtained and analysed the results it produced. EO wrote the first draft of the paper. IC conducted the manual LR of the use case and IC and EO identified the relevant papers. EO, IC, Ath and AM helped write the paper and EO, AM, OK, AC and IC reviewed the paper.

## Acknowlegments

Ms. Mafalda Vieira Burri, the librarian from the library of the University of Geneva who helped define the search queries, Mr. Alexander Temerev, doctoral student at the Institute of Global Health, who helped write part of the Python code on parallelising the APIs.

## Abbreviations

AEHI: Acute and early hiv infection
API: Application programming interface
DBCV: Density-based clustering validation
HDBSCAN: Hierarchical density-based spatial clustering of applications with noise
HIV: Human immunodeficiency virus
k-NN: k-nearest neighbours
LR: Literature review
NLP: Natural language processing
PacMAP: Pairwise controlled manifold approximation
TF-IDF: Term frequency - Inverse document frequency
UML: Unsupervised machine learning
WSS: Work saved over sampling

## Supplementary Material

### Query for Embase

(‘acute hiv’:ab,ti OR ‘early hiv’:ab,ti OR ‘primary hiv’:ab,ti OR (‘window period’:ab,ti AND ‘human immunodeficiency virus’:ab,ti)) AND (‘africa south of the sahara’/exp OR ‘sub-saharan africa’:ab,ti OR ‘subsaharan africa’:ab,ti OR ‘africa south of the sahara’:ab,ti OR angola:ab,ti OR benin:ab,ti OR botswana:ab,ti OR ‘burkina faso’:ab,ti OR burundi:ab,ti OR cameroon:ab,ti OR ‘cape verde’:ab,ti OR ‘central africa’:ab,ti OR ‘central african republic’:ab,ti OR chad:ab,ti OR comoros:ab,ti OR congo:ab,ti OR ‘cote divoire’:ab,ti OR ‘democratic republic congo’:ab,ti OR djibouti:ab,ti OR ‘equatorial guinea’:ab,ti OR eritrea:ab,ti OR eswatini:ab,ti OR ethiopia:ab,ti OR gabon:ab,ti OR gambia:ab,ti OR ghana:ab,ti OR guinea:ab,ti OR ‘guinea-bissau’:ab,ti OR kenya:ab,ti OR lesotho:ab,ti OR liberia:ab,ti OR madagascar:ab,ti OR malawi:ab,ti OR mali:ab,ti OR mayotte:ab,ti OR mozambique:ab,ti OR namibia:ab,ti OR niger:ab,ti OR nigeria:ab,ti OR rwanda:ab,ti OR sahel:ab,ti OR ‘sao tome and principe’:ab,ti OR senegal:ab,ti OR ‘sierra leone’:ab,ti OR somalia:ab,ti OR ‘south africa’:ab,ti OR ‘south sudan’:ab,ti OR sudan:ab,ti OR tanzania:ab,ti OR togo:ab,ti OR uganda:ab,ti OR zambia:ab,ti OR zimbabwe:ab,ti)

### Query for Web of Science

TS=((“acute hiv” OR “early hiv” OR “primary hiv” OR (“window period” AND “human immunodeficiency virus”)) AND (“africa south of the sahara”/exp OR “sub-saharan africa” OR “subsaharan africa” OR “africa south of the sahara” OR angola OR benin OR botswana OR”burkina faso” OR burundi OR cameroon OR “cape verde” OR “central africa” OR “central african republic” OR chad OR comoros OR congo OR “cote divoire” OR “democratic republic congo” OR djibouti OR “equatorial guinea” OR eritrea OR eswatini OR ethiopia OR gabon OR gambia OR ghana OR guinea OR “guinea-bissau” OR kenya OR lesotho OR liberia OR madagascar OR malawi OR mali OR mayotte OR mozambique OR namibia OR niger OR nigeria OR rwanda OR sahel OR “sao tome and principe” OR senegal OR “sierra leone” OR somalia OR “south africa” OR “south sudan” OR sudan OR tanzania OR togo OR uganda OR zambia OR zimbabwe))

**Table S1.**
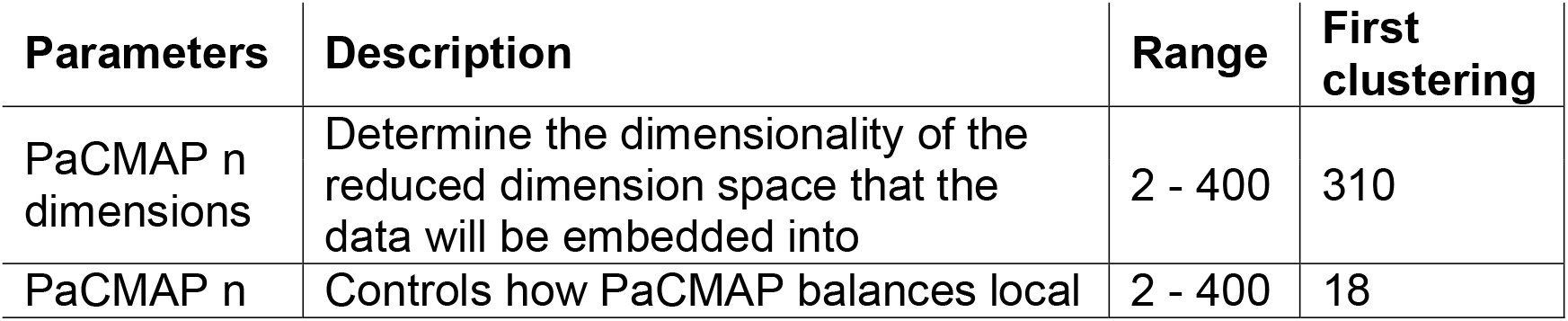
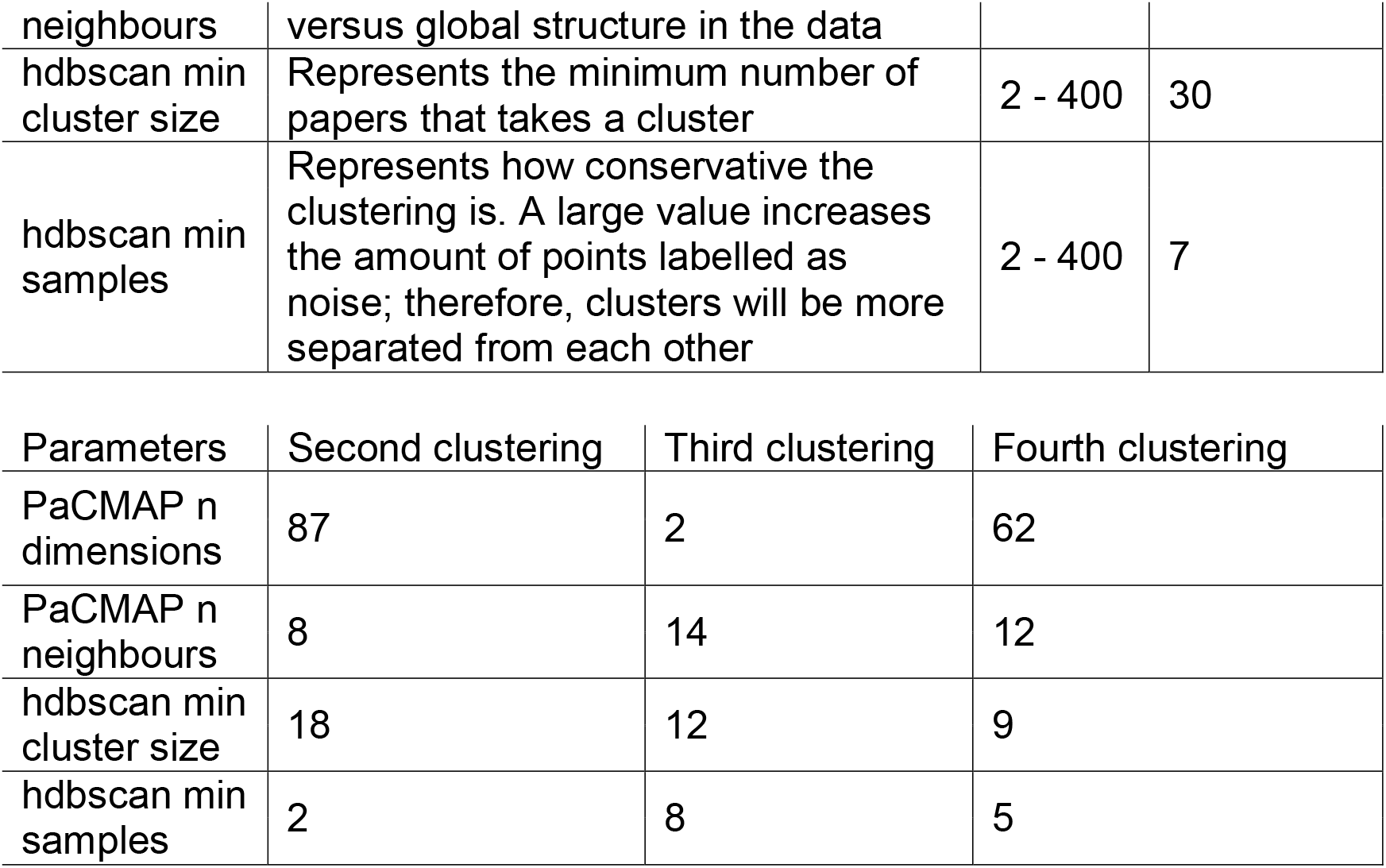
Hyperparameters definition, range and values for each clustering

